# Anterior Segment Optical Coherence Tomography in Pediatric Ocular Pathology: Imaging Study of 115 eyes

**DOI:** 10.1101/2022.01.31.22270061

**Authors:** Sana Nadeem

## Abstract

**Background:** To assess the pediatric anterior segment characteristics in ocular pathology, using spectral domain optical coherence tomography (SD-OCT).

**Methods:** Our case series follows 115 eyes of 78 children aged 2–17 years of age with anterior segment pathology in an academic facility. A thorough eye examination and investigations were performed for each child. The anterior segment optical coherence tomography (AS-OCT) analysis was done using the Optopol Revo 80^®^ high resolution SD-OCT (840 nm, axial resolution of 5 µm) using an imaging adapter. All pathological features visible on imaging were observed, studied, tabulated, and analyzed.

**Results:** 115 eyes of 78 children with anterior segment pathology were imaged. The average age was 11.84 years, with 44 males and 34 females. The primary clinical diagnosis was *cataract* (congenital and acquired) in 40 (34.8%) eyes, followed by *corneal disease* (congenital, inflammatory, and traumatic) in 28 (24.3%) eyes, *glaucoma* (juvenile and secondary) in 18 (15.7%) and *trauma* in 15 (13%) eyes. Systemic diseases were associated in 20.9%. The commonest imaging pathology observed was *lens opacification* (any morphology/location) in 43 (37.4%), *increased reflectivity of the cornea* in 31 (28.2*%), corneal stromal thinning* in 34 (29.6%), increased corneal thickness in 28 (24.3%), *shallow anterior chamber* in 17 (14.8%), and *cells in anterior chamber* in18 (15.7%) eyes, along with a multitude of other findings.

**Conclusions:** Our study demonstrates that anterior segment optical coherence tomography is a useful non-contact technique used without sedation for the detailed anatomic and pathologic assessment, imaging, diagnosis, and monitoring of pediatric ocular diseases.

## 1. INTRODUCTION

Optical coherence tomography (OCT) is a non-invasive, rapid investigative imaging modality which provides intricate micron resolution three-dimensional images, for the diagnosis and monitoring of both anterior and posterior segment disorders. It utilizes infrared light reflected by the ocular structures to create high resolution cross sectional images. Its utility in very small children is limited because of non-cooperation especially in very young children, the need for a chin rest and focusing on the part of the children. However, it is a non-contact procedure, and is generally well tolerated. Anterior segment OCT (AS-OCT) is a rapidly emerging technology which provides high resolution images of the structures constituting the anterior segment, especially the cornea, ocular surface, angle of anterior chamber, iris, lens and any pathology pertaining to the anterior segment can be viewed in extreme detail, provided the media is sufficiently clear. It can also be used as a measuring tool for corneal thickness, anterior and posterior chamber depths, anterior chamber diameter, chamber angle configuration and even iris and lens thicknesses. It can help immensely in the diagnosis and monitoring of anterior segment disorders by providing useful information about pediatric ocular anterior segment disease. ^1-5^

The rationale of our study is to investigate the AS-OCT in pediatric eyes presenting with any pathology of the anterior segment, to image the diseases and to describe the features visualized by this emerging technique.

## 2. MATERIAL and METHODS

A total of 78 cooperative children, aged 2 to 17 years, presenting to our outpatient and inpatient department with any anterior segment disorder were included in our study. Non-cooperative children and those with severe intellectual disabilities were excluded, due to inability to mount their head on the chin and forehead rest of the machine and also inability to follow to fixation target. This is a 15-month study carried out from 31^st^ August, 2020 till 7^th^ December, 2021 in the department of Ophthalmology of Fauji Foundation Hospital, which is a tertiary care teaching hospital of the Foundation University Islamabad, Pakistan. Permission from the ERC (Ethical review committee) of Foundation University Medical College was taken previously, which is in concordance with the declaration of Helsinki. Parental informed consent was taken prior to evaluation.

Each child was thoroughly evaluated with visual acuity estimation done with the Snellen chart or the Lea chart, depending upon the age of the patient; routine refraction (cycloplegic or automated) and a complete anterior and posterior segment evaluation was done with the slit lamp, intraocular pressure estimation and B-scan ultrasonography was done in case of media opacity, as the case may be.

The anterior segment OCT evaluation was done with the Optopol Revo 80^®^ high resolution SD-OCT (840 nm, transverse resolution 12 µm; axial resolution 5 µm) which provides 80,000 A scans per second. It provides a scan depth of 2.4 mm and a range of 3-16 mm and 3D, Radial, B-scan, Raster, Cross scans can be done for every patient. Images were obtained at the horizontal (0° and 180°), vertical (90° and 270°) and oblique meridians (135° and 315°; 45° and 225°) as well. The examination was carried out in the sitting position for young children, either in the parent’s lap or standing position if very small. It is a non-contact device, with no need for any anesthesia. The child and parents were explained the procedure and the child asked to place his/her head on the chin and forehead mount of the OCT machine with the additional imaging adapter mounted on the lens which is provided with the device to allow wide and detailed scans of anterior segment. The child was asked to open the eye wide to prevent shadowing from the eyelids and eyelashes and look into the adapter and follow the internal fixation target (green cross) which is in the default mode. The eye was aligned and good quality images were captured.

High resolution imaging of the cornea, its thickness, abnormal reflectivity or other pathology like scar depth, corneal burn depth, and tears was obtained; the anterior chamber depth and configuration; a detailed assessment of the anterior chamber angle structures including iris root, angle recess, Schlemm’s canal, scleral spur, in some cases anterior ciliary body and anterior trabecular meshwork were done; lens pathology, absence, precise cataract morphology and location, anterior and posterior capsules as well as posterior capsular presence or absence or lenticonus, intraocular lens position, and even Elschnig’s pearls, anterior PHPV could be studied; and iris pathology including flattening, tears, holes and iridodialyses were examined as well. Anterior chamber cells, exudate and hyphema could be visualized as well as synechiae of the iris with the lens and cornea. Trabeculectomy bleb and fistula was also examined in one child as well as post-operative pseudophakic children or trauma cases in order to monitor intraocular lens (IOL) position and surgical integrity. was Conjunctival tumor characteristics and scleral pathology including scleritis and trauma were also studied by asking the child to move the eye into the desired position or by using an external fixation target (red light); to view the desired area. High resolution images were obtained and studied. In the event of poor vision as in cases of total cataracts, phthisis bulbi, total corneal opacification or severe trauma, the child was simply asked to gaze into the adapter and good quality images were obtained still. The AS-OCT images obtained by our device are in grayscale or monochrome representing the backscattered light intensity in a cross-sectional scheme. The grayscale is directly related to the tissue reflectivity. False colours may be used to delineate boundaries of various structures like the corneal layers but may create artificial boundaries. Surgical planning was also done after visualizing the extent of trauma, cataract and posterior capsular status. Measurements of the corneal scar, lens opacity, iris thickness, conjunctival tumors and angle of anterior chamber was also done with extreme detail for future monitoring with the line measurement tool and angle measurement tool of the OCT, respectively.

The patients were classified according to the primary clinical diagnosis. [Table 1] Associated systemic diseases were tabulated as well. The structural pathology on AS-OCT was also documented and analyzed and tabulated. [Table 2]

**Table 1:** Primary clinical diagnosis for each eye (More than one diagnosis/feature may be present in the same eye)

| Clinical Diagnosis | Frequency (Percent) |
| --- | --- |
| *Cataract* | 40 (34.8) |
| • Congenital / Developmental Cataract | 30 (26.1) |
| • Secondary Cataract (trauma, drugs, uveitis) | 18 (15.7) |
| *Corneal Disease* | 28 (24.3) |
| • Corneal scar/ tear | 8 (7) |
| • Corneal opacification | 6 (5.2) |
| • Keratoconus/hydrops | 8 (7.0) |
| • Cornea plana | 4 (3.5) |
| • Microcornea | 2 (1.7) |
| • Band keratopathy | 2 (1.7) |
| • Peter's anomaly | 1 (0.9) |
| • OMMP<sup>α</sup> | 1 (0.9) |
| *Glaucoma* | 18 (15.7) |
| • JOAG<sup>β</sup> | 14 (12.2) |
| • Secondary glaucoma | 3 (2.6) |
| • Neovascular glaucoma | 1 (0.9) |
| *Trauma* | 15 (13) |
| • Blunt trauma | 9 (7.8) |
| • Penetrating trauma | 5 (4.3) |
| • Thermal trauma (burn) | 1 (0.9) |
| Iris flattening/tear | 4 (3.5) |
| *Pseudophakia* | 4 (3.5) |
| *Conjunctival tumors* | 3 (2.6) |
| • Conjunctival Melanocytic Nevus | 2 (1.7) |
| • Corneo-scleral dermoid | 1 (0.9) |
| *Ectopia lentis* | 3 (2.6) |
| *Nodular anterior scleritis* | 1 (0.9) |
| *Anterior Uveitis* | 1 (0.9) |
| *Phthisis bulbi* | 1 (0.9) |
| *PHPV<sup>γ</sup>* | 1 (0.9) |
| *Persistent pupillary membrane* | 1 (0.9) |
| *Associated systemic disease* | 24 (20.9) |
| • HLH<sup>Ω</sup> | 2 (1.7) |
| • JIA<sup>∞</sup> | 4 (3.5) |
| • Nephrotic syndrome | 2 (1.7) |
| • Deaf mutism | 2 (1.7) |
| • Intellectual disability | 2 (1.7) |
| • MMP<sup>α</sup> | 1 (0.9) |
| • GERD<sup>π</sup> & Cleft Palate | 1 (0.9) |
| • Depression & Anxiety (Floxetine) | 2 (1.7) |
| • Diabetes mellitus Type 2 | 2 (1.7) |
| • Marfan syndrome | 2 (1.7) |
| • Epidermolysis bullosa | 2 (1.7) |
| • Down's syndrome | 2 (1.7) |

α = Ocular/mucous membrane pemphigoid β = Juvenile open angle glaucoma γ Persistent hyperplastic primary vitreous Ω = Hemophagocytic lymphohistiocytosis ∞ = Juvenile idiopathic arthritis π = Gastroesophageal reflux disease

**Table 2:** Anterior segment pathology as seen on AS-OCT imaging

| **AS-OCT PATHOLOGY** | **FREQUENCY (PERCENT)** |
| --- | --- |
| **CORNEAL DISEASE** |  |
| Increased corneal epithelial/stromal/endothelial reflectivity | 34 (29.6) |
| Increased corneal thickness | 28 (24.3) |
| Corneal bullae | 5 (4.3) |
| Corneal stromal thinning | 31 (27) |
| Corneal tear |  |
| • Partial thickness/lamellar laceration | 1 (0.9) |
| • Total thickness/self-sealing laceration | 5 (4.3) |
| • Total thickness with iris prolapse | 1 (0.9) |
| **ANTERIOR CHAMBER** |  |
| Cells in anterior chamber | 18 (15.7) |
| Fibrinous exudate in anterior chamber | 3 (2.6) |
| Shallow anterior chamber | 17 (14.8) |
| Anterior displacement of iris-lens diaphragm | 2 (1.7) |
| Vitreous in anterior chamber | 2 (1.7) |
| Emulsified silicon oil in anterior chamber | 1 (0.9) |
| **CATARACT MORPHOLOGY/OTHER LENS PATHOLOGY** |  |
| • Total or diffuse (mild or dense) or with cortical clefts | 17 (14.8) |
| • Lamellar, nuclear and punctate | 2 (1.7) |
| • Sutural + Coronary | 2 (1.7) |
| • Sutural | 1 (0.9) |
| • Posterior Polar | 1 (0.9) |
| • Primary aphakia | 1 (0.9) |
| • Sutural + PSCO<sup>€</sup> | 2 (1.7) |
| • Punctate + nuclear + sutural + PSCO<sup>€</sup> | 2 (1.7) |
| • Lens rupture with total, soft cataract | 1 (0.9) |
| • Anterior Polar/Pyramidal + Sutural | 1 (0.9) |
| • PSCO<sup>€</sup> | 12 (10.4) |
| • ASCO<sup>£</sup> | 1 (0.9) |
| • PCO<sup>¥</sup> | 6 (5.2) |
| • Punctate | 4 (3.5) |
| • ASCO<sup>£</sup> + PSCO<sup>€</sup> + Punctate | 1 (0.9) |
| • ASCO<sup>£</sup> + PSCO<sup>€</sup> | 4 (3.5) |
| • Nuclear Lamellar Pulverulent | 6 (5.2) |
| Lens opacification (any opacity) | 43 (37.4) |
| Lens subluxation | 3 (2.6) |
| Anterior capsular fibrosis | 14 (12.2) |
| Pseudophakia | 9 (7.8) |
| Posterior capsular fragility/absence | 1 (0.9) |
| Posterior lenticonus | 1 (0.9) |
| Primary aphakia/aphakia other | 3 (2.6) |
| **ANGLE** |  |
| Narrow angle | 15 (13) |
| Wide open angle | 100 (87) |
| Abnormal angle tissue | 1 (0.9) |
| **IRIS PATHOLOGY** |  |
| Iris strands attached to endothelium | 4 (3.5) |
| Iris flattening/other iris damage | 15 (13.6) |
| Iris holes/tears | 1 (0.9) |
| Iridodialysis | 2 (1.7) |
| Posterior synechiae | 13 (11.3) |
| Pupillary membrane | 1 (0.9) |
| **CONJUNCTIVAL PATHOLOGY** |  |
| Conjunctival flap | 1 (0.9) |
| Conjunctival tumor characteristics |  |
| • Dome shaped mass | 3 (2.6) |
| • Intrinsic cysts | 2 (1.7) |
| • Posterior shadowing | 3 (2.6) |
| • Heterogeneous mass | 3 (2.6) |
| • Suprascleral fluid | 2 (1.7) |
| Subconjunctival silicon oil | 1 (0.9) |
| Trabeculectomy bleb- diffuse | 1 (0.9) |
| **SCLERAL PATHOLOGY** |  |
| • Scleral tear (partial thickness) | 1 (0.9) |
| • Scleral tear (full thickness) | 1 (0.9) |
| Heterogeneous hyporeflexivity | 1 (0.9) |
| Scleral thickening | 1 (0.9) |
| **VITREOUS PATHOLOGY** |  |
| Vitreous cells | 2 (1.7) |

€ = Posterior subcapsular opacity ¥ = Posterior capsular opacification £ = Anterior subcapsular opacity

SPSS version 20 was used to analyze the data. Frequencies and percentages were calculated for age, gender, eye laterality, clinical diagnosis, associated systemic disease and anterior segment pathologies/findings on AS-OCT imaging.

## 3. RESULTS

Our pediatric cohort included 115 eyes of 78 children [44 (38.3%) males and 34 (29.6%) females] presenting to us in a 15-month period. The predominant primary clinical diagnosis was *cataract* in 40 (34.8%) eyes,, This included congenital and developmental cataract in 26.1% eyes and secondary to trauma, uveitis and steroid or fluoxetine usage in 15.7%. *Corneal disease* came second with 28 (24.3%) eyes including corneal scars, keratoconus, and congenital anomalies like cornea plana, microcornea and Peter’s anomaly. *Glaucoma* came in third in 18 (15.7%) with the majority being JOAG, steroid induced in the rest and one case of neovascular glaucoma (NVG) in advanced retinoblastoma. *Trauma* accounted for a total of 15 (13%) eyes, with blunt trauma in the majority. Conjunctival tumors and ectopia lentis accounted for 3 (2.6%) eyes each. Congenital diseases accounted for the majority of our cohort. [Table 1] 24 (20.9%) eyes had a positive systemic association tabulated in Table 1.

### 3.1. Uses of AS-OCT in Corneal and Ocular surface diseases

AS-OCT has been invaluable in studying corneal and conjunctival disease. Increased corneal reflectivity was observed in 34 (29.6%) eyes with varying pathologies ranging from corneal tears, corneal scars at varying depths [8 (7%)], corneal opacification in 5.2%, calcium deposition in band keratopathy [2 (1.7%)] and Peter’s anomaly [1(0.9%)]. Corneal thickness was increased in 28 (24.3%) eyes, mostly with corneal edema, manifesting also with bullae [5(4.3%)]. Corneal stromal thinning was observed in 31 (27%) eyes with mostly scars and keratoconus. Conjunctival flap was seen in one case of ocular mucus membrane pemphigoid (OMMP) which was treated surgically with a Gunderson flap for a previous perforation. Subconjunctival silicon oil was observed in one eye with secondary glaucoma with a previous pars plana vitrectomy and silicon oil tamponade. [Figure 1]

**Figure 1:**
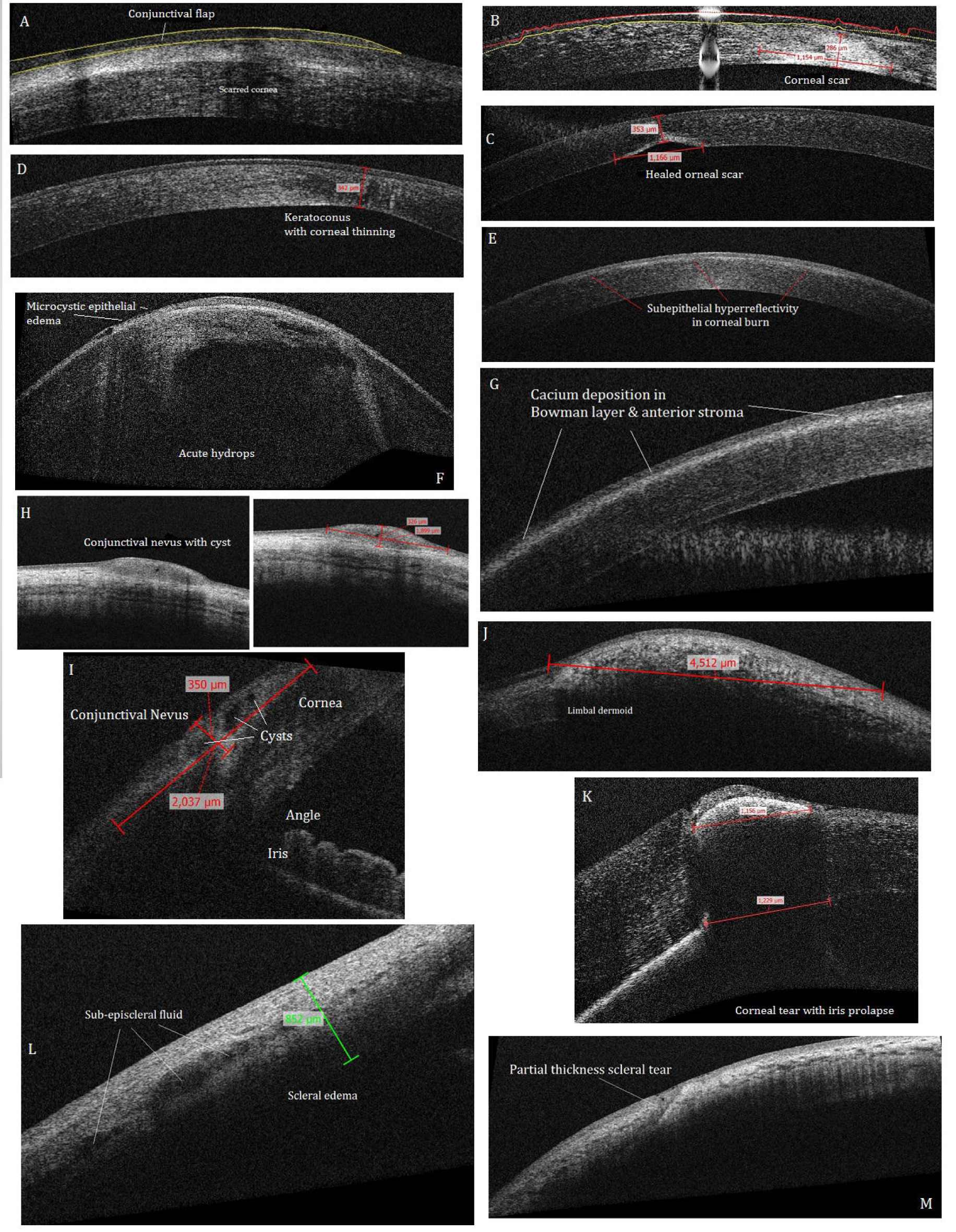
Various corneal, conjunctival and scleral pathologies seen on AS-OCT

### 3.2. Uses of AS-OCT in Anterior Chamber and angle evaluation

The anterior chamber configuration can be assessed with the wide radial scan and also the full range radial scan. Cells in uveitis can be clearly observed, so can emulsified silicon oil but these cannot be distinguished on images [Fig 3]. Narrowing of the angle was observed in 15 (13%) of the eyes with mostly trauma, but also tumors, and ectopia lentis. Detailed AC angle evaluation was done in all glaucoma cases and all of the JOAG cases [14 (12.2%)] had a wide open angle and abnormal angle tissue was observed in one eye only, which was not visible on gonioscopy [Fig 3L]. The Schlemm’s canal was visible in all cases of JOAG and secondary steroid induced glaucoma. NVG secondary to retinoblastoma had a narrow angle and anterior displacement of the iris and lens diaphragm.

### 3.3. Uses of AS-OCT in Anterior Segment Tumors

Conjunctival nevi [2(1.7%)] were visualized to be heterogeneous, diffuse or dome shaped masses with intrinsic cysts and posterior shadowing. Conjunctival limbal dermoid [1(0.9%)] was imaged as a heterogeneous mass with posterior shadowing. [Figure H-J]

### 3.4. Uses of AS-OCT in Lens disorders

Any degree of lens opacification was observed in 43(37.4%) of the eyes with most common being total cataract [17(14.8%)], posterior subcapsular opacity (PSCO) in 12 (10.4%) and numerous other morphologies, mostly mixed. Posterior capsular opacification was observed clearly in 6 (5.2%) with one case of very large Elschnig pearls pushing the iris forwards. Anterior capsular fibrosis was another common observation in 14 (12.2%), and posterior capsular fragility and absence was clearly delineated prior to surgery, posterior lenticonus was seen as a conical protrusion backwards and primary aphakia was seen as absence of the lens behind the iris with a clear space in front of the persistent hyperplastic primary vitreous (PHPV). Congenital ectopia lentis in a case of Marfan’s syndrome showed bilateral nasal subuxation of the lenses. [Figure 1]

### 3.5. Uses of AS-OCT in Iris disorders

Iris flattening [15(13.6%)] was observed in most cases of JOAG, secondary glaucoma and trauma. Posterior synechiae were visualized in 13 (11.3%) eyes, mostly with trauma and uveitis. Iris tears, iridodialysis, and persistent pupillary membrane were observed in one case each. Peter’s anomaly was visualized as iris strands attached to a posterior corneal defect in one eye. Similar iris anterior synechiae were seen in two additional eyes of penetrating trauma.

### 3.6. Uses of AS-OCT in Trauma

A detailed evaluation of traumatized eyes was possible with extent of the corneal or scleral tears demarcated; traumatic uveitis, exudate, synechiae and cataracts including lens rupture were observed as well. Corneal tears were classified into lamellar or partial thickness [1(0.9%)], total thickness, self sealing [5(4.3%)] and total thickness with iris prolapse [1(0.9%)]. Scleral tears were similarly diagnosed and classified into partial thickness and full thickness 1(0.9%)] each [Fig 1]. Iridodialysis and iris tears were observed as well 1(0.9%)]. Traumatic cataract included total white, lens rupture and ASCO or PSCO. Traumatic lens subluxation was observed in one eye [Fig 2].

**Figure 2:**
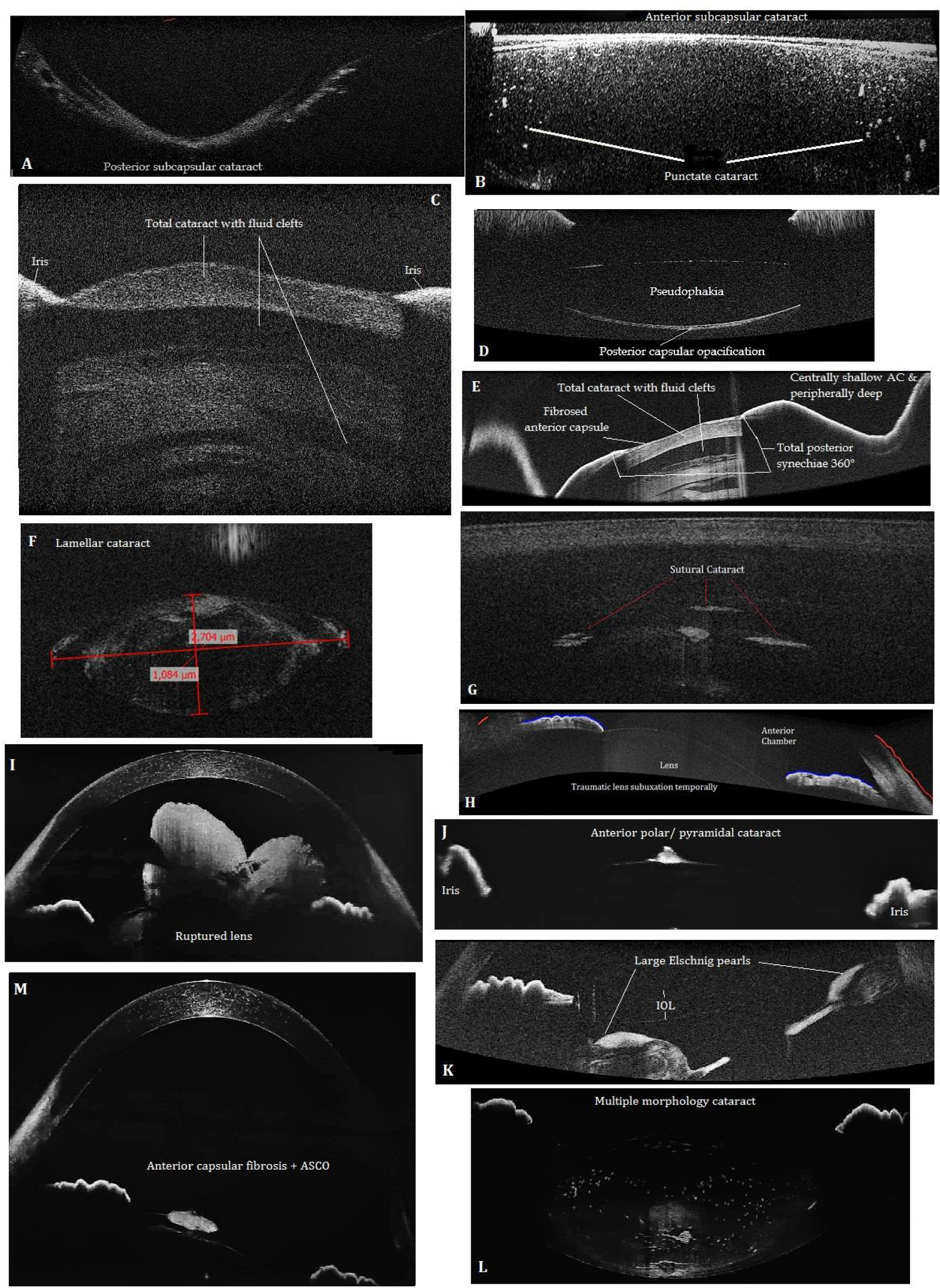
The various types of cataract and lens disorders seen with AS-OCT (self-explanatory)

**Figure 3:**
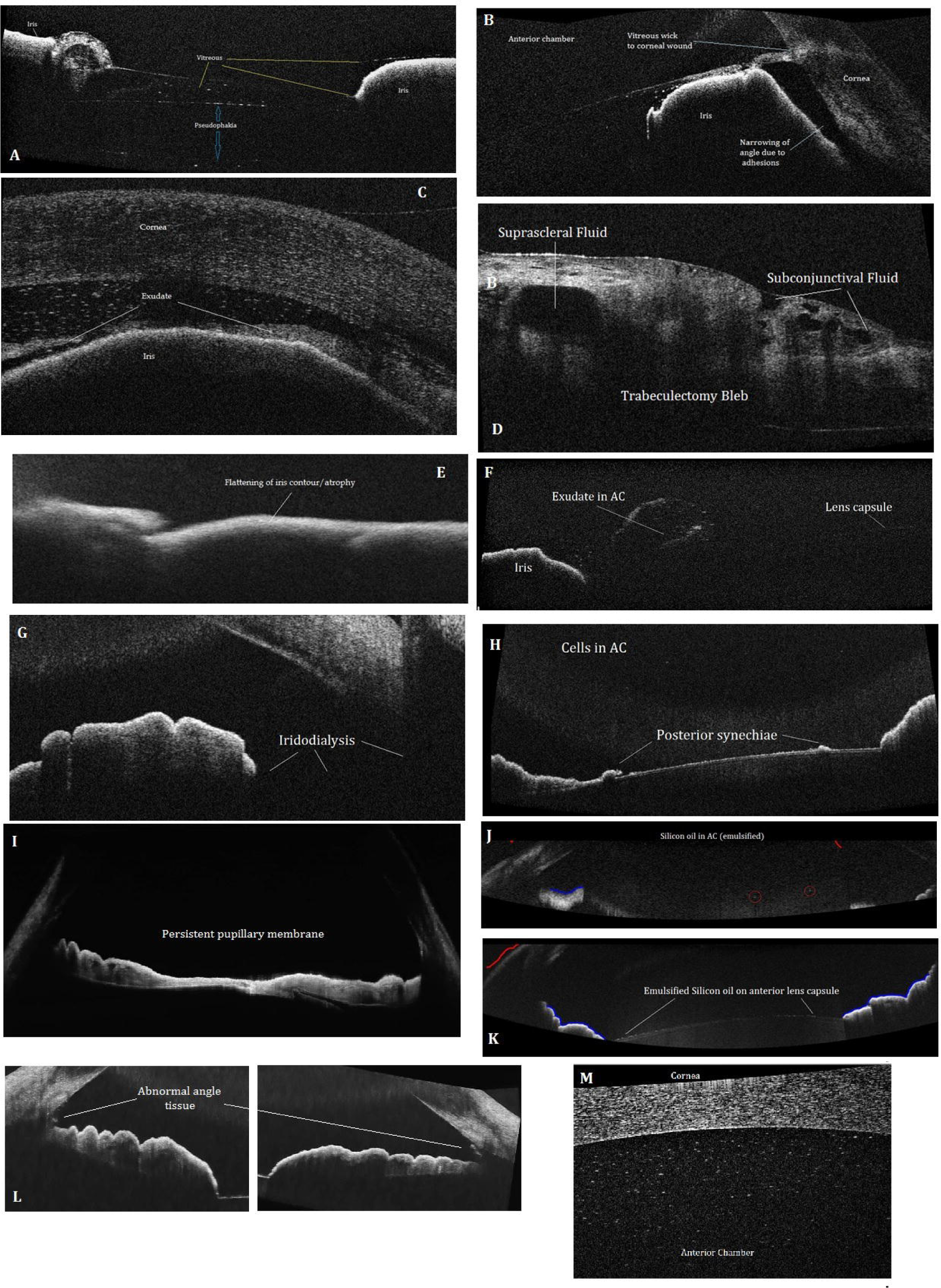
A. Vitreous wick over IOL B. Narrow angle and vitreous wick to corneal wound C. Shallow anterior chamber with serosanguinous exudate and cells/microhyphema D. A functioning trabeculectomy bleb E. Iris flattening in blunt trauma F. Fibrinous exudate over lens G. Iridodialysis H. Remnants of posterior synechiae on anterior lens capsule I. Persistent pupillary membrane J. Emulsified silicon oil in AC K. Emulsified silicon oil over anterior lens capsule L. Abnormal angle tissue in one eye of JOAG M. Cells in anterior chamber

### 3.7. Uses of AS-OCT in Scleral diseases

Scleritis was confirmed in one case with scleral thickening, heterogeneous hyporeflectivity and subepiscleral fluid pockets observed. [Fig 1L]

### 3.8. Uses of AS-OCT in Surgical planning and evaluation

Planning of corneo-scleral repairs, cataract surgery and even post-operative evaluation of repairs, IOL position, trabeculectomy fistula and bleb evaluation was also done.

## 4. DISCUSSION

OCT has found a leading position in research and ophthalmic practice after its introduction in the 1990s. It is based upon the principle of *optical reflectometry*^1^, utilizing the measurement of back-scattered light through transparent or semi-transparent biological tissues. SD-OCT uses the principle of *low-coherence interferometry* to acquire in vivo, in situ, cross sectional imaging of the anterior or posterior segments with histological resolution.^6^ AS-OCT, since its advent in 1994, has been subsequently been utilized to provide an *‘optical biopsy’*^7^ and to evaluate pediatric ocular diseases, by only a handful of researchers so far. The applications of this technology are manifold in children including pachymetry, ocular surface disease, conjunctival and corneoscleral pathology including ectatic disorders, opacity and scarring, anterior segment tumors, pediatric glaucoma, biometry, cataract, uveitis, anterior segment dysgenesis and evaluation of surgical procedures on these structures, including keratoplasty and trabeculectomy. ^2-14^

Cauduro et al.^9^ have beautifully described the usage of AS-OCT in pediatric ophthalmology in 26 eyes of 19 children with various anterior segment diseases in their article, and have labelled it feasible in clarifying diagnosis, and surgical planning and follow up. Majander^10^ et al. have utilized this technique with extensive benefit in pediatric eyes of 7 patients with varying causes of congenital corneal opacities. We have imaged both congenital and acquired corneal pathology and found it to be extremely useful. Bradfield^12^ et al. have compared angle structures in their unique subset of 8 children with glaucoma of any type versus healthy controls and found developmental angle anomalies including absent or anomalous Schlemm’s canal, using hand-held OCT. We found only one case of abnormal angle tissue nasally and temporally in one eye of JOAG. Pilat^13^ et al have utilized hand-held OCT in 6 children with anterior segment dysgenesis to study its features. Anjos et al.^15^ have studied anterior segment parameters in 17 children with primary congenital glaucoma (PCG) and compared them with normal children and have described this as a useful modality in the follow up and prognosis of this disease. They found iris hypoplasia, similar to Pilat et al.^16^, who examined 22 patients with PCG and found iris flattening and thin collarette zone along with high iris insertion. Gupta et al.^17^ have reported abnormal angle tissue, hyperreflective membranes and absence of the Schlemm canal in some of their PCG and JOAG patients. We have found iris flattening in the majority of JOAG patients and abnormal, hyperreflective angle tissue in one case. Qiu^18^ et al. in their study on myopic children have found the anterior segment to be elongated. Bhatti et al.^19^ have elaborated a clinical classification of pediatric uveitis based on AS-OCT imaging. Cells can be easily observed and quantified with AS-OCT as well. We have even been able to image emulsified silicon oil droplets in the AC, subconjunctivally and adherent to the anterior lens capsule. Bajwa et al.^20^ have studied aniridia with this technique in 43 patients, some of them were children. Ahmed^21^ et al. have utilized this technique in 9 children with mucopolysaccharidoses and found it useful to predict glaucoma risk, and also observed crowded anterior segments and thick corneas in these. Sridhar^22^ et al. have described its use for corneal deposits, dystrophies, keratitis and contact lens practice amongst other uses. We have also found it to be very useful in delineating corneal pathology, edema, calcium deposits in band keratopathy and scar demarcation. Chan et al.^23^ have reported use of this technology to image eyes with posterior polar cataracts and grade them and posterior capsular deficiency, thus to identify risk of posterior capsular rent, and rendering it useful for better patient diagnosis, preparation and management. We also used this to evaluate the status of posterior capsule prior to congenital cataract surgery and in trauma assessment and planning. Posterior capsular defects, fragility and posterior lenticonus can be visualized with clarity. Lepska et al.^24^ have imaged 74 Polish children with AS-OCT and have termed it invaluable in the diagnosis, planning and operations in anterior segment pediatric disorders. They used general anesthesia for evaluation in the majority of infants and children in their cohort. For our study, we did not require any sedation and anesthesia, in as young as a 2-year-old child, because mostly our cohort comprised of older and cooperative children. Strengths of our study are that it is the largest study on pediatric cohort utilizing anterior segment OCT and also the first of its kind in our country. We have added a variety of anterior segment disorders to elaborate the pathological findings on AS-OCT imaging.

Limitations of our study are that we failed to include infants, smaller children and non-cooperative ones. The hand-held OCT would be more informative for such cases; as ours was a conventional OCT with a head and chin rest which required considerable cooperation on the part of the patient, and excusion of infants and smaller children.

Our SD-OCT at 840 nm has provided detailed anterior segment imaging. However, higher resolution swept source OCT (SS-OCT) machines at 1310 nm would provide even more detailed anterior segment imaging, especially of the deeper angle structures, like trabecular meshwork and ciliary body.^7,14, 25^

## 5. CONCLUSION

Our study reflects that anterior segment OCT is a useful non-contact technique without sedation for detailed anatomic and pathologic assessment, imaging, diagnosis and monitoring of pediatric ocular diseases.

## Data Availability

The data for this study is available from Open Science Framework with the DOI 10.17605/OSF.IO/8EANC and link: https://osf.io/8eanc/?view_only=5106ee4478d441f1ad60a68a2b72259d

## Abbreviations

AC: anterior chamber
ASCO: Anterior subcapsular opacity
AS-OCT: Anterior segment optical coherence tomography
GERD: Gastroesophageal reflux disease
JIA: Juvenile idiopathic arthritis
JOAG: Juvenile open angle glaucoma
HLH: Hemophagocytic lymphohistiocytosis
IOL: intraocular lens
NVG: neovascular glaucoma
OCT: optical coherence tomography
OMMP: Ocular/mucous membrane pemphigoid
PCG: primary congenital glaucoma
PCO: Posterior capsular opacification
PHPV: Persistent hyperplastic primary vitreous
PSCO: Posterior subcapsular opacity
SD-OCT: spectral domain OCT
nm: nanometer
µm: micrometer

